# SARS-CoV-2 genomic surveillance in Rwanda: Introductions and local transmission of the B.1.617.2 (Delta) variant of concern

**DOI:** 10.1101/2022.05.31.22275802

**Authors:** Yvan Butera, Samuel L. Hong, Muhammed Semakula, Nena Bollen, Verity Hill, Áine Niamh O’Toole, Barney I. Potter, Dieudonné Mutangana, Reuben Sindayiheba, Robert Rutayisire, Maria Artesi, Vincent Bours, Nadine Rujeni, Simon Dellicour, Keith Durkin, Leon Mutesa, Guy Baele

## Abstract

The emergence of the SARS-CoV-2 Delta variant of concern (lineage B.1.617.2) in late 2020 resulted in a new wave of infections in many countries across the world, where it often became the dominant lineage in a relatively short amount of time. We here report on a novel genomic surveillance effort in Rwanda in the time period from June to September 2021, leading to 201 SARS-CoV-2 genomes being generated, the majority of which were identified as the Delta variant of concern. We show that in Rwanda, the Delta variant almost completely replaced the previously dominant A.23.1 and B.1.351 (Beta) lineages in a matter of weeks, and led to a tripling of the total number of COVID-19 infections and COVID-19-related fatalities over the course of only three months. We estimate that Delta in Rwanda had an average growth rate advantage of 0.034 (95% CI 0.025-0.045) per day over A.23.1, and of 0.022 (95% CI 0.012-0.032) over B.1.351. Phylogenetic analysis reveals the presence of at least seven local Delta transmission clusters, with two of these clusters occurring close to the border with the Democratic Republic of the Congo, and another cluster close to the border with Tanzania. A smaller Delta cluster of infections also appeared close to the border with Uganda, illustrating the importance of monitoring cross-border traffic to limit the spread between Rwanda and its neighboring countries. We discuss our findings against a background of increased vaccination efforts in Rwanda, and also discuss a number of breakthrough infections identified during our study. Concluding, our study has added an important collection of data to the available genomes for the Eastern Africa region, with the number of Delta infections close to the border with neighboring countries highlighting the need to further strengthen genomic surveillance in the region to obtain a better understanding of the impact of border crossings on lowering the epidemic curve in Rwanda.

## Introduction

The world experienced a surge of COVID-19 cases with the emergence of the Deltavariant (lineage B.1.617.2) in late 2020 (Saito et al., 2022), resulting in scarcity of oxygen supplies and hospitals being overwhelmed in many countries (Vaidyanathan, 2021). In May 2021, the Delta variant first observed in India quickly became the dominant lineage worldwide. As of 11 July 2021, 89 countries across six continents had reported the presence of Delta (*Cov-Lineages*, n.d.), which was of particular concern for African countries due to vaccine inequality and the potentially devastating effects on unvaccinated individuals. Rwanda experienced its third wave of COVID-19 in the beginning of June 2021 as demonstrated by a substantial increase in the test positivity rate and the number of daily new infections (Figure 1). During the month of May 2021, the test positivity rate was on average 1.4%, whereas in June 2021, it was 5.4% to become 11.4% in July 2021 and 4.7% in August 2021. On May 31, 2021, the total number of people who had been diagnosed with COVID-19 in Rwanda since the first case on March 14, 2020, equalled 26,963; twelve weeks later this number more than tripled, with a total number of 88,682 as of September 2, 2021. The number of COVID-19 hospitalizations related to COVID-19 went from 46 on May 31, 2021 to 2,687 on September 2, 2021. Since the beginning of the pandemic up to May 2021, Rwanda has registered 353 deaths. On September 2, 2021, mortality related to COVID-19 was still limited to 1,105 people (RBC, n.d.). This low overall mortality rate has been noted throughout the pandemic, with previous reports showing that Rwanda only observed 133 COVID-19 deaths in the ten months after its first COVID-19 case (Musanabaganwa et al., 2021). Africa in general observed a lower mortality ratio compared to the rest of the world, with 168 deaths per million people, whereas the ratio is 402, 2160 and 2739 per million in South-East Asia, Europe, and Northern America, respectively (*WHO Coronavirus (COVID-19) Dashboard*, n.d.). There is a rationale that the lower mortality rate in Africa was linked to the demographic characteristics of the continent (Lawal, 2021), where the median age is 19.7 years, while it is 32 in Asia, 42.5 in Europe and 38.6 in Northern America (*World Population Prospects - Population Division - United Nations*, 2019). Only 0.3 percent of the population is above 80 years of age in sub-saharan Africa compared to 1.2 % in Asia, 3.8% in Europe and 3.2 % in Northern America (*World Population Prospects - Population Division - United Nations*, 2019). However, the fact that sub-Saharan nations were reporting lower case numbers and fewer COVID-19 deaths than might be expected has been linked to a deficit of testing and strained medical infrastructure (Gill et al., 2022). On the other hand, a study conducted in West Africa on blood of people who had been exposed to Ebola and Lassa Fever showed that, compared to healthy blood donors and COVID-19 patients in the United States of America, they had more exposures to coronaviruses with epitopes that cross react with SARS-CoV-2, and therefore induce cross-reactive immunity that may protect against death (Borrega et al., 2021).

**Figure 1.**
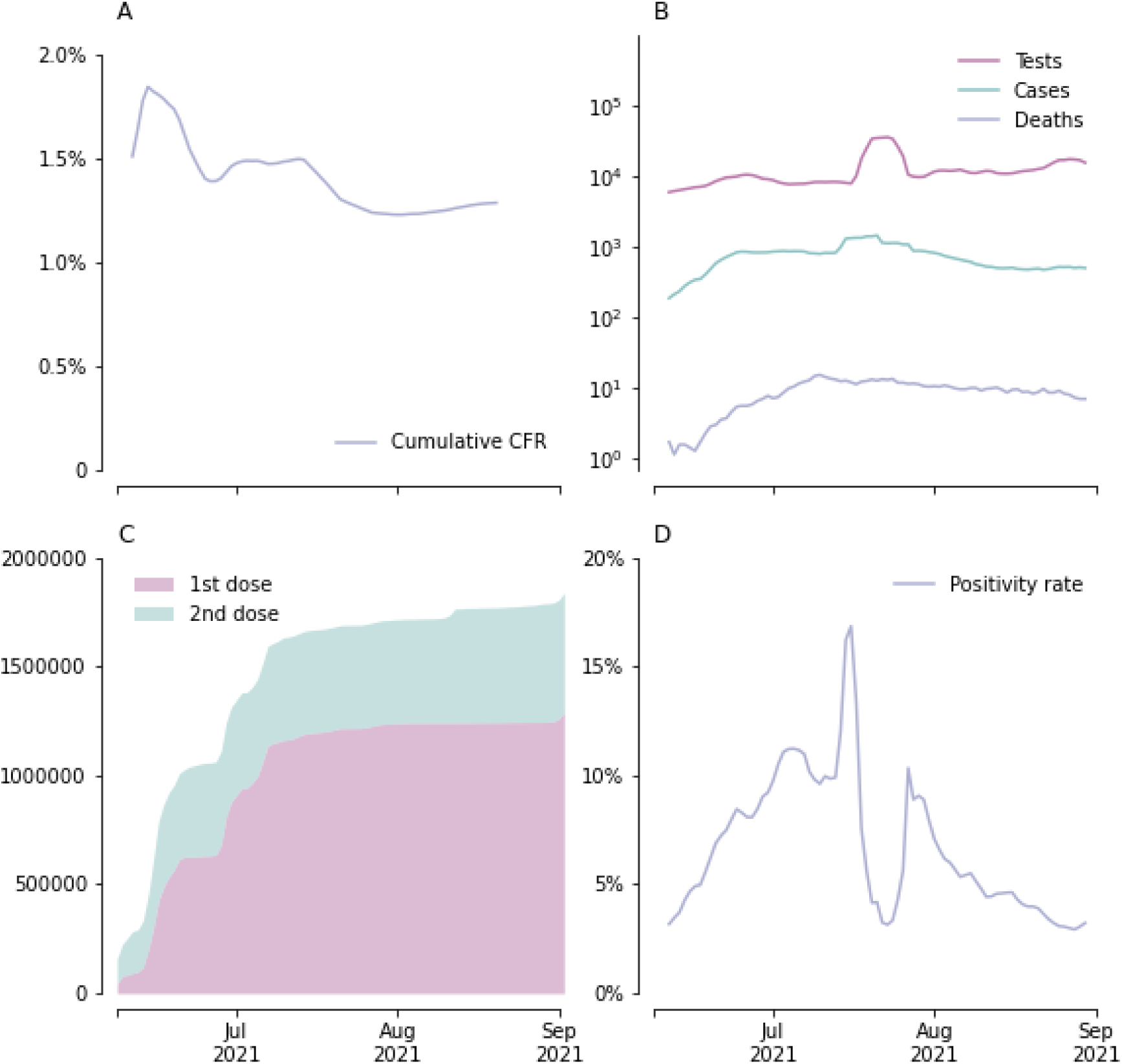
A) Cumulative case fatality ratio (CFR) 7-day rolling average; B) Daily tests, cases and deaths, 7-day rolling average; C) Cumulative number of vaccinated individuals; D) Testing positivity rate 7-day rolling average. An abrupt drop in both CFR and testing positivity rate is observed late July 2021. This can be explained by the considerable increase in the number of tests performed during the same time period.

In light of the increasing positivity rate in June, the government of Rwanda tightened preventive measures, where a curfew was established countrywide at 9 PM. Public offices, restaurants and cafés operated at 30% capacity, whereas social gatherings occurring in homes were prohibited. On July 14, the capital City of Kigali and eight other districts entered into a full lockdown amid a continued surge in cases. Movements of people were not allowed and schools were closed except for critical services like healthcare and groceries shopping. On August 11, the lockdown was lifted and a curfew was instated at 8pm. Public and private offices were allowed to resume functioning at 50% capacity, and restaurants at 30%.

## Vaccination Status

Rwanda is among the first of the African countries that had started COVID-19 vaccination campaigns early in March 2021. The Government of Rwanda applied for different vaccines, including Pfizer-BioNTech, Moderna, and AstraZeneca vaccines among others, and had also submitted all required documents to COVAX, a framework aiming to ensure equitable access and fair allocation of COVID-19 health products. Rwanda purchased additional vaccines to fast track vaccination rollout. Vaccination has been shown to be an essential aspect to limit the numbers of infections and deaths. For example, a study in the United States showed that during October-November 2021, in a period of Delta predominance and Omicron emergence, unvaccinated persons had 13.9 and 53.2 times the risks for infection and COVID-19-associated death, respectively, compared with fully vaccinated persons who received booster doses, and 4.0 and 12.7 times the risks compared with fully vaccinated persons without booster doses (Johnson et al., 2022).

Though currently Rwanda aims to vaccinate all inhabitants 18 years and older, initially the primary beneficiaries were people at high risk, such as health professionals, people with comorbidities, and the elderly of 65 years and older. For sustainability, Rwanda is working with BioNTech to establish vaccine manufacture within the country, an initiative that aims to also serve other African countries (*IFC, Government of Rwanda Partner to Develop Vaccine Manufacturing Capacity in Rwanda*, 2021). The country established supply chain infrastructures including procuring ultra-cold freezers and containers, storage and proper distribution mechanisms of vaccines in each of the four provinces of Rwanda.

Rwanda adopted vaccination as one of many preventive measures against the COVID-19 pandemic. The country’s goal to vaccinate 70% of its population (approximately 7.8 million people) by mid-2022 has been attained (*Rwanda Biomedical Centre*, 2020). Although more than 6 billion vaccine doses were administered globally by September 2021, there is a clear discrepancy in distribution with less than 2% of vaccines having been delivered to Africa (Massinga Loembé & Nkengasong, 2021). This despite the fact that the average vaccination acceptance rate is higher in low and middle income countries (LMICs) than in high income countries (HICs) such as the United States and Russia (Solís Arce et al., 2021). The solution to such stark vaccine access inequity might arise from recent developments such as the partnership that African countries are rolling out with BioNTech that will see platforms <u>that will</u> allow for mRNA vaccine production in bulk being delivered in the second half of 2022 in Rwanda, Senegal, and South Africa (*Africa Is Bringing Vaccine Manufacturing Home*, 2022, “mRNA Made in Africa,” 2022).

As of August 2021, the vaccination campaign has resulted in over 1.6 million inhabitants having received the first dose, which corresponds to 21% of targeted people, and 12% of targeted people are fully vaccinated. The vaccination campaign focused on high risk areas, therefore in the capital City of Kigali 93% of targeted people have received their first dose and 46% are fully vaccinated (Figure 2). An observational test-negative case–control study (Lopez Bernal et al., 2021) regarding vaccine effectiveness against symptomatic disease showed that the Delta variant more often led to disease symptoms compared to the Alpha / B.1.1.7 variant, both for one and two vaccine doses of either Pfizer-BioNTech or AstraZeneca, although the difference in vaccine effectiveness was much smaller among persons who had received the second dose of vaccine. These data, showing the importance of being vaccinated, came on top of concerns of researchers about the Delta variant - owing to its increased transmissibility which could overwhelm healthcare systems - spreading beyond urban centers to rural regions that were largely spared in earlier waves of the pandemic and have less access to testing, healthcare and vaccines (Mallapaty, 2021) (see Figure 3 for major hospitals across Rwanda).

**Figure 2.**
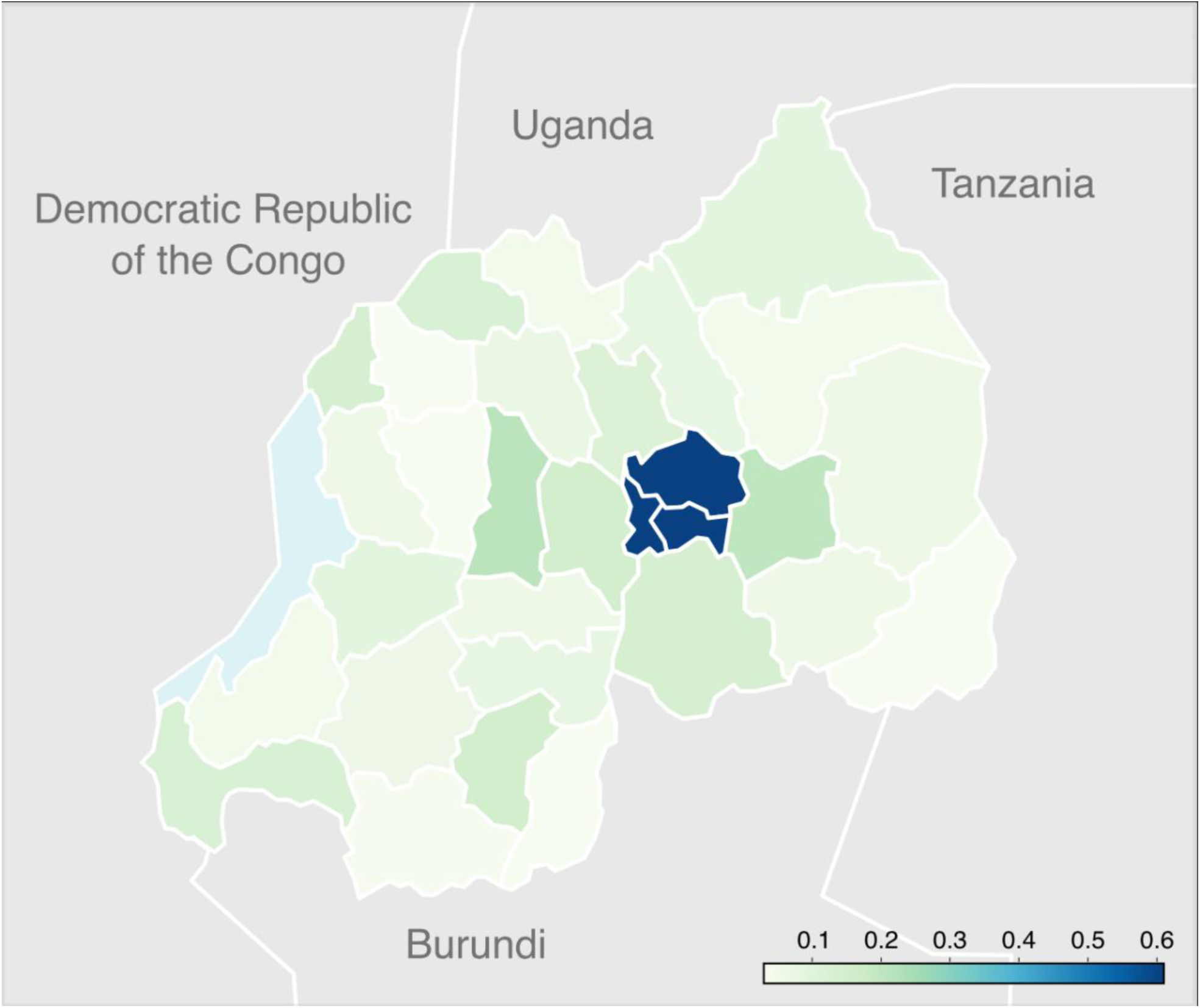
Cumulative vaccination coverage in the provinces of Rwanda up to September 2, 2021. The map shows the proportion of people having received at least one dose. The highest vaccination coverage is found in the province of Kigali, which has a noticeably higher vaccination coverage than the rest of the country (60%, compared to less than 20% for most other provinces).

**Figure 3.**
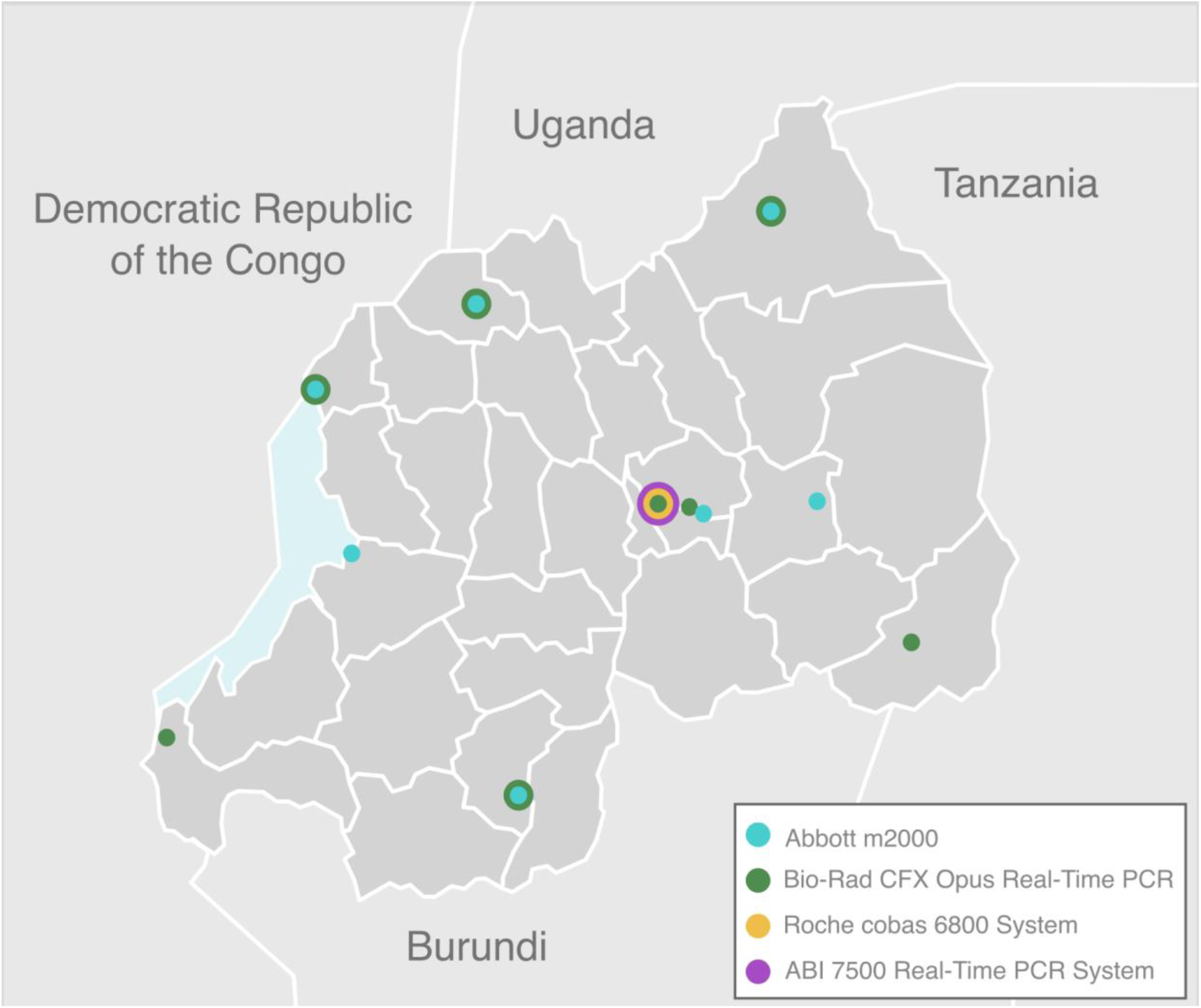
RT-PCR based testing sites across Rwanda. There are 17 testing platforms distributed across 10 teaching and referral hospitals, and the National Reference Laboratory in Kigali, Rwanda. There are two types of automated SARS-CoV-2 testing platforms used: the Abbott m2000 and the Roche 68000. The manual platforms are: Bio-Rad CFX Opus Real-Time PCR and ABI 7500 Real-Time PCR system.

## Results

We performed a sample collection from individuals residing in different provinces of Rwanda, from 17 testing platforms distributed across Rwanda that conduct RT-PCR based testing for SARS-CoV-2 (Figure 3). Out of the 201 Rwandan samples collected and sequenced as part of this study, 161 (80.1%) were Delta / B.1.617.2, with a further 5.0% belonging to the different AY.* pango lineages (2 AY.11, 1 AY.23, 1 AY.24, 1 AY.30 and 5 AY.7.1). Nine genomes (4.5%) were classified as pango lineage A.23.1, which was the dominant lineage at the end of our previous study in January-February 2021 (Butera et al., 2021). At that time, we had picked up the first cases of the Beta variant of concern (lineage B.1.351), which accounted for 13 (6.5%) of the genomes collected in this new study. Finally, we found single occurrences of the B.1.214.2, B.1.214.4 and B.1.351.3 pango lineages. Of note, our data collection initiative covered a large part of Rwanda, including the capital region as well as regions bordering all neighboring countries, with only a few provinces from which no samples were available for this study (Figure 4). We show the gradual progression over time of the different lineages identified during our study period in Supplementary Figure S1. At the end of our study period, in September 2021, we only sampled genomes from pango lineages Delta B.1.617.2 and its various AY.* sublineages.

**Figure 4.**
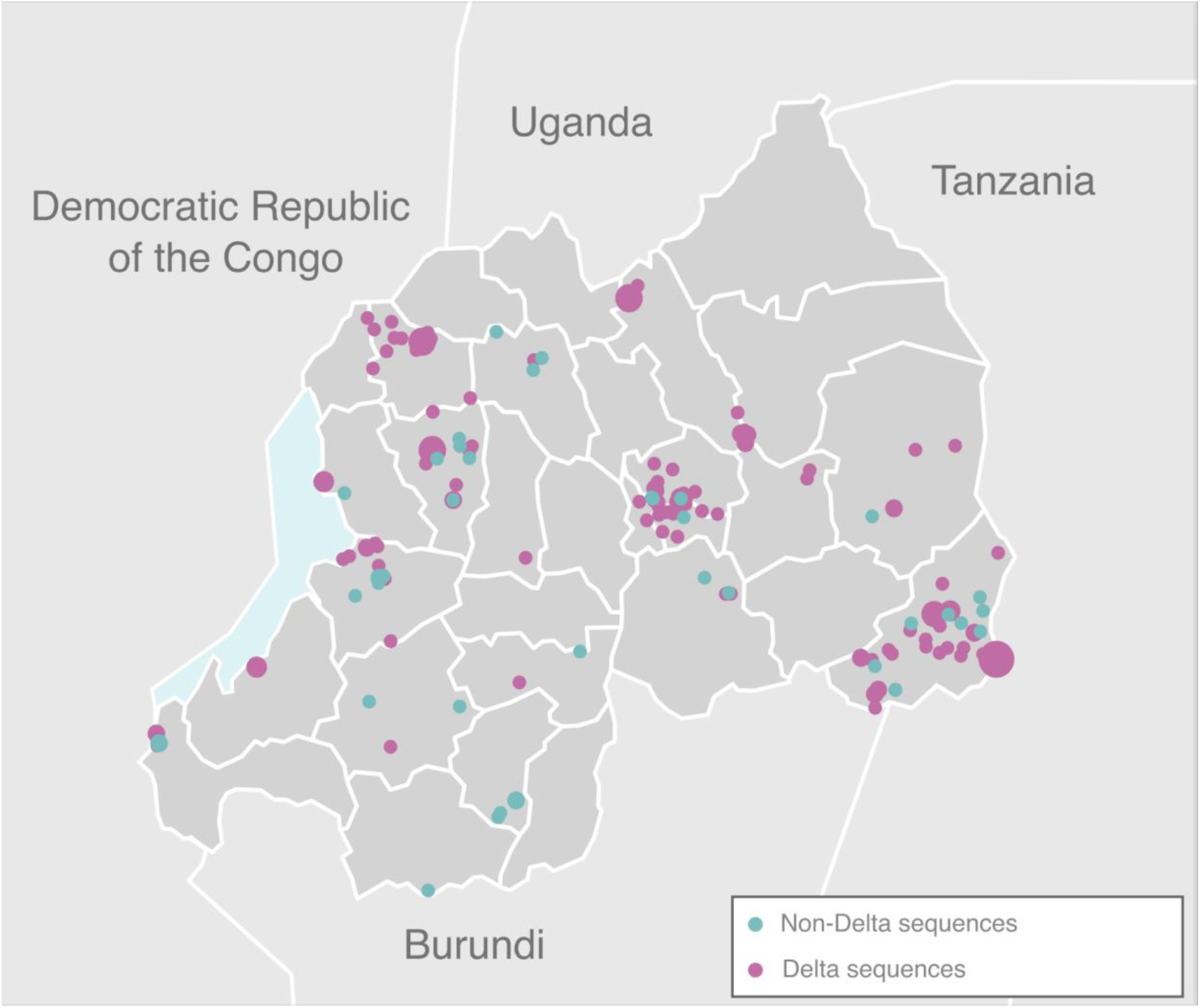
Geographic distribution of the collected genomes in this study. Delta / B.1.617.2 sequences (*n* = 159) are shown in pink and all other lineages (*n* = 39) in blue. Three sequences did not have geographic location up to village level and are hence not shown.

Starting March 2021, Delta rapidly displaced the most abundant lineages A.23.1 and B.1.351, and became the dominant variant circulating nationally by June 2021 (Figure 5A & 5B). This swift replacement was also observed elsewhere in Africa, including in countries that are geographically close to Rwanda (Figure 6). This pattern was also observed for example in Benin, where Delta increased from accounting for 3.4% of sequenced positive samples in May 2021, to 37.9% in June and 63.6% in July 2021 (Yadouleton et al., 2022). We estimate that Delta had an average growth advantage of 0.034 (95% CI: 0.025-0.045) per day compared to A.23.1 and of 0.022 (95% CI: 0.012-0.032) compared to B.1.351 (Figures 8A & 8B). Assuming that the variants have the same generation time of 5.2 days (Ganyani et al., 2020), this corresponds to a growth advantage relative to A.23.1 of 20% (95% CI: 13-26%) per generation of viral transmission, and of 12% (95% CI: 7-17%) over Beta (B.1.351). During the same time period, we notice a slight increase in the relative abundance of Alpha (B.1.1.7), accounting for up to 4% of the proportion of variants circulating at the time during the last days of May. Despite the overwhelming dominance of Delta during the same time period, our modeling results show only a mean growth rate advantage of 9% (95%: CI 1-17%) with respect to Alpha. We estimate the largest increase in competitive advantage when comparing Delta to B.1.380. In this case, our results show a growth rate difference of 0.14 (95% CI: 0.06-0.23), which translates to an increase in effective reproductive number of 110% (95% CI: 33-233%). Such an increase in growth advantage is not surprising, since B.1.380 was in the process of becoming extinct in the country by the time Delta was starting to widely circulate. Lastly, when compared to other minority variants, we notice that Delta has a growth rate increase of 0.33 (95%: CI 0.02 - 0.04) and hence a growth advantage of 19% (95% CI: 12-26%). Overall, our modeled estimates of growth rate advantage for Delta in Rwanda appear to be lower than those previously reported using global data (Campbell et al., 2021).

**Figure 5.**
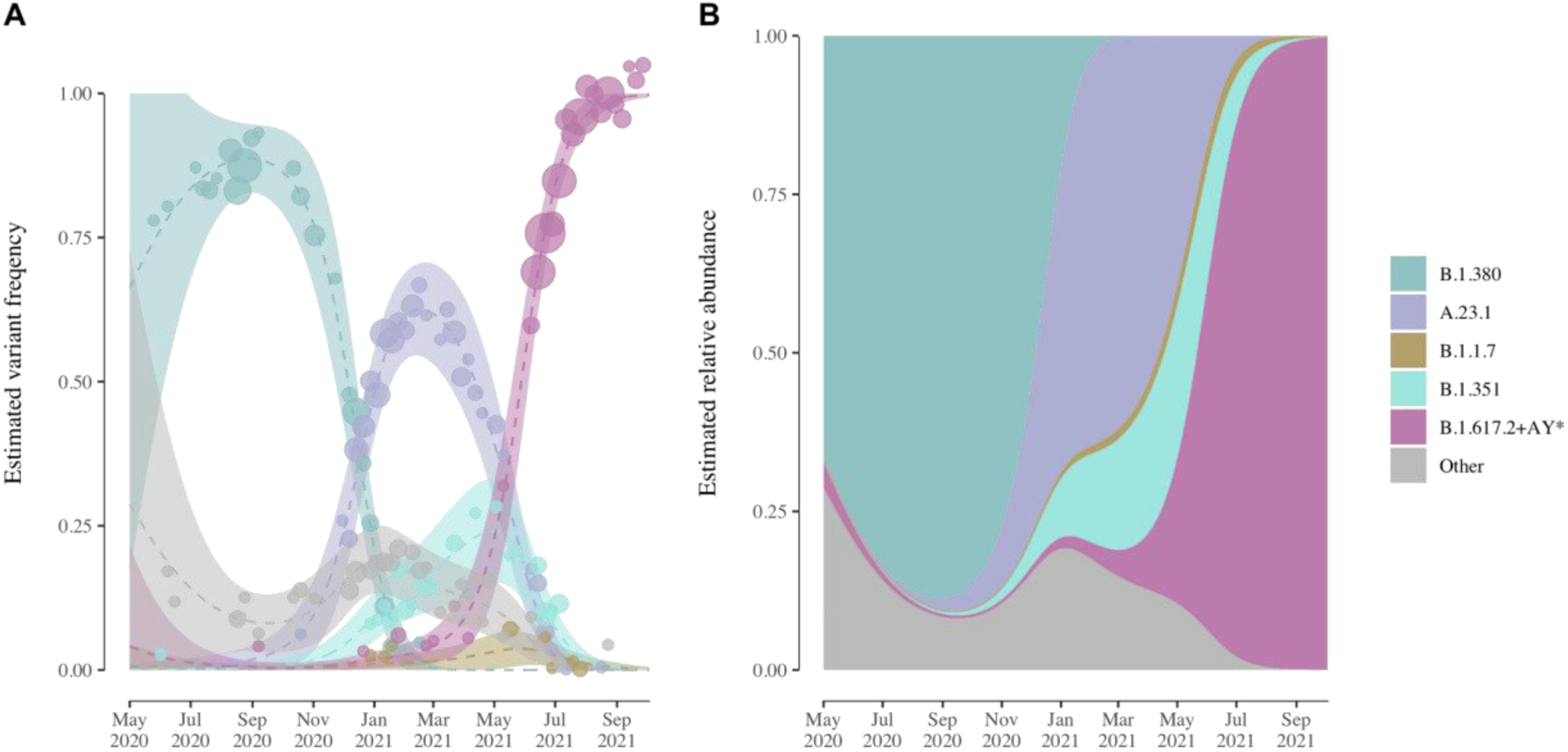
A) Modeled estimates of the frequency of each lineage of interest. Only the previously dominant lineages B.1.380 and A.23.1 and variants of concern (VOC) B.1.1.7 (Alpha), B.1.351 (Beta) and B.1.617.2+AY.* (Delta and sublineages) are represented by individual colors. Other minority non-VOC lineages are collapsed under the ‘Other’ category. Dashed lines represent mean estimates and shaded regions 95% confidence intervals. Dots correspond to the sequences collected each week, with the size corresponding to the number of sequences collected; B) Muller plot showing the population dynamics of SARS-CoV-2 lineages in Rwanda over time. Estimates of the proportion of circulating variants show the replacement of lineage B.1.380 by lineages A.23.1 and B.1.351, and the subsequent rapid replacement by Delta.

**Figure 6.**
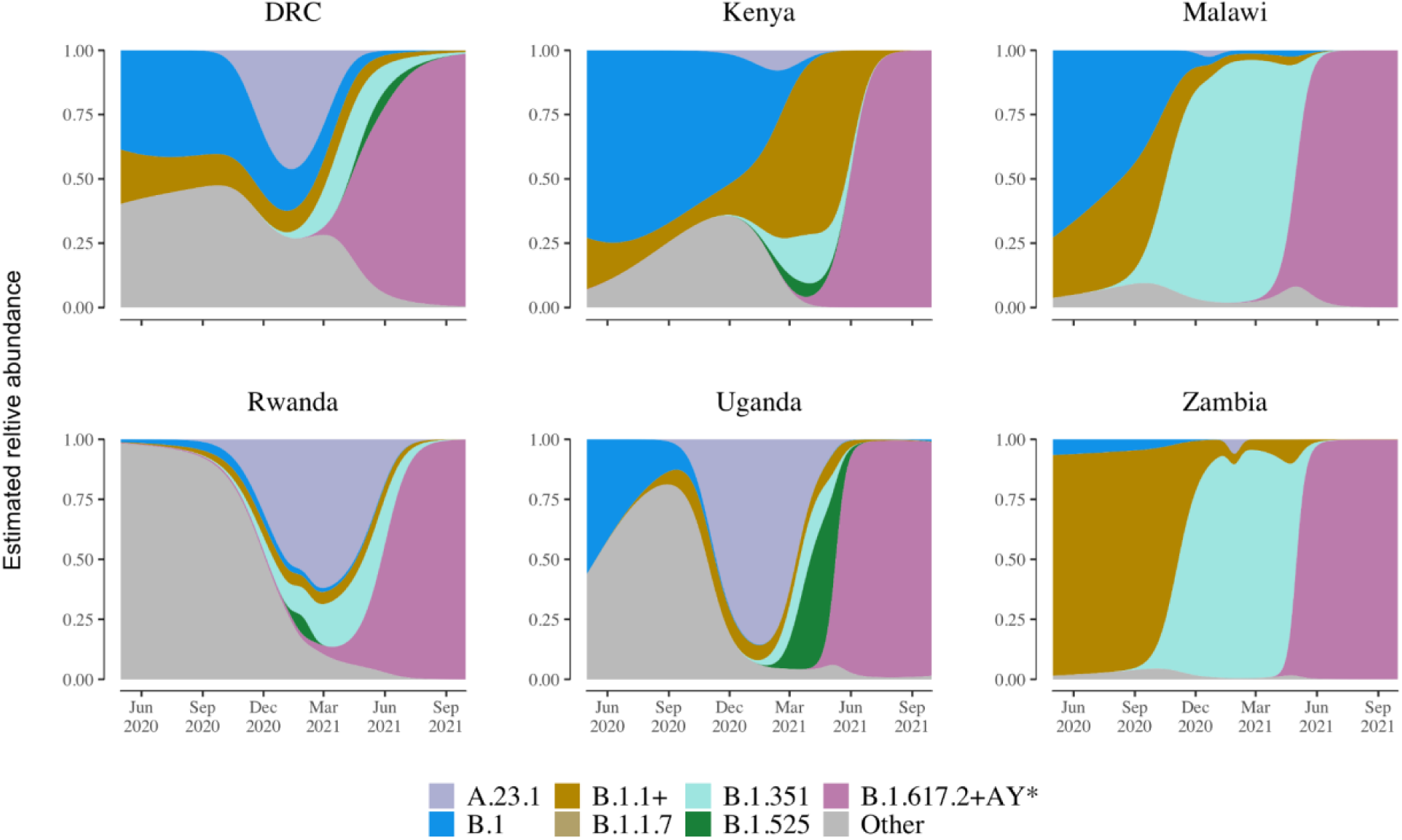
Relative abundance of SARS-CoV-2 variants of concern in Rwanda and other African countries in the region. Only geographically close countries with over 200 sequences collected between June 2020 and September 2021 are included (source: GISAID). Individual colors were assigned to lineages depending on their VOC status and prevalence in Rwanda. B.1.1+ refers to lineage B.1.1 and all non-VOC descendants. Other minority variants are collapsed under the category of Other. Although prior to April 2021, the pattern of dominant lineages differs by country, by July 2021 the Delta variant (B.1.617.2+AY.*) had become the dominant lineage in the region.

**Figure 7.**
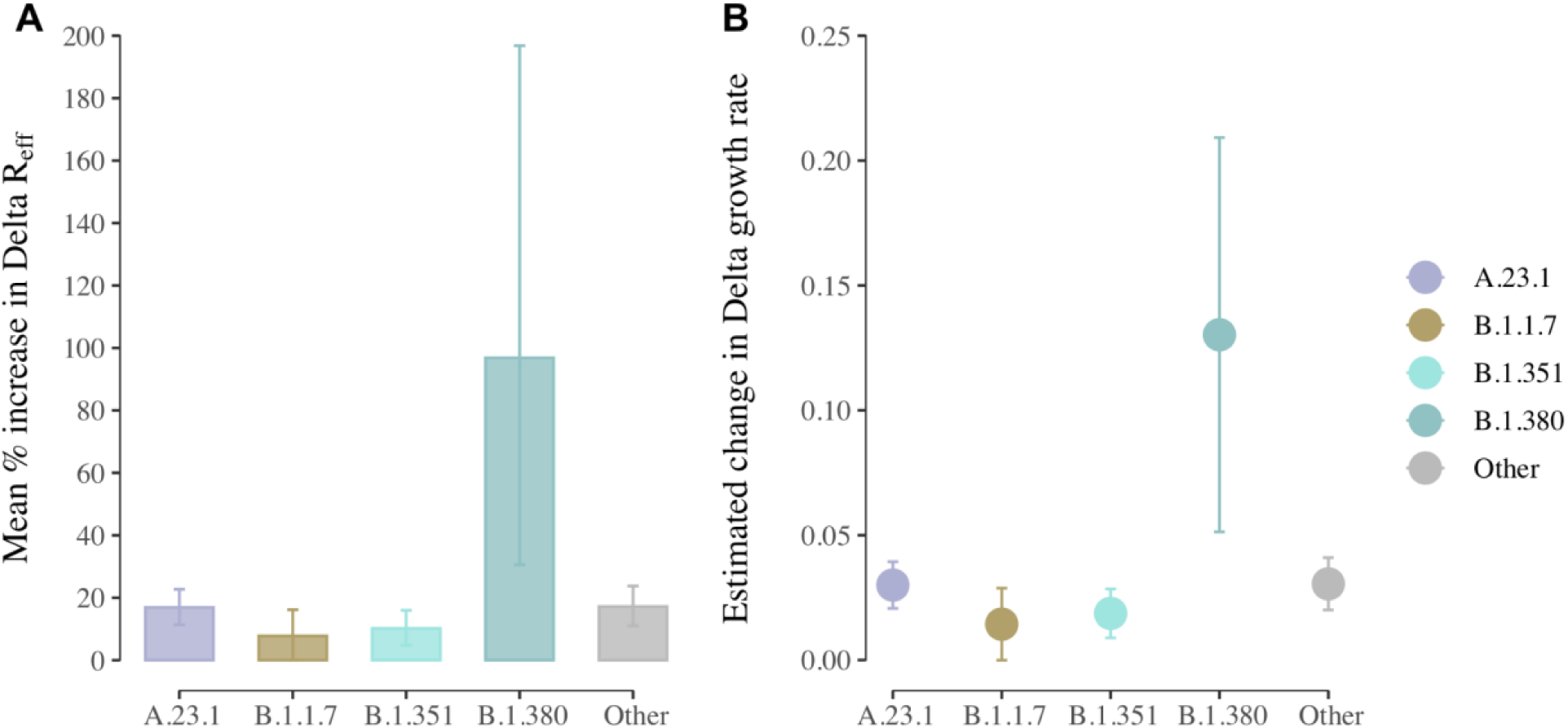
A) Transmission advantage of Delta relative to the other lineages of interest. Transmission advantage is defined as the percentage increase in the effective reproduction number of a lineage relative to another. Thus in the case of a transmission advantage of 20% relative to A.23.1, the effective reproductive number of Delta is 20% higher to that of A.23.1. The estimates shown here were obtained from a multinomial logistic spline model, and an assumed equal generation time across all variants of 5.2 days; B) Estimated change in growth rate of Delta relative to all other lineages. The plot depicts the mean and 95% confidence intervals of the difference in Malthusian growth rates between Delta and all other variants as estimated from a multinomial logistic spline model. Given the non-linear variation in growth rates across time, the estimates were averaged across the time period between September 10, 2020 and September 30, 2021.

**Figure 8.**
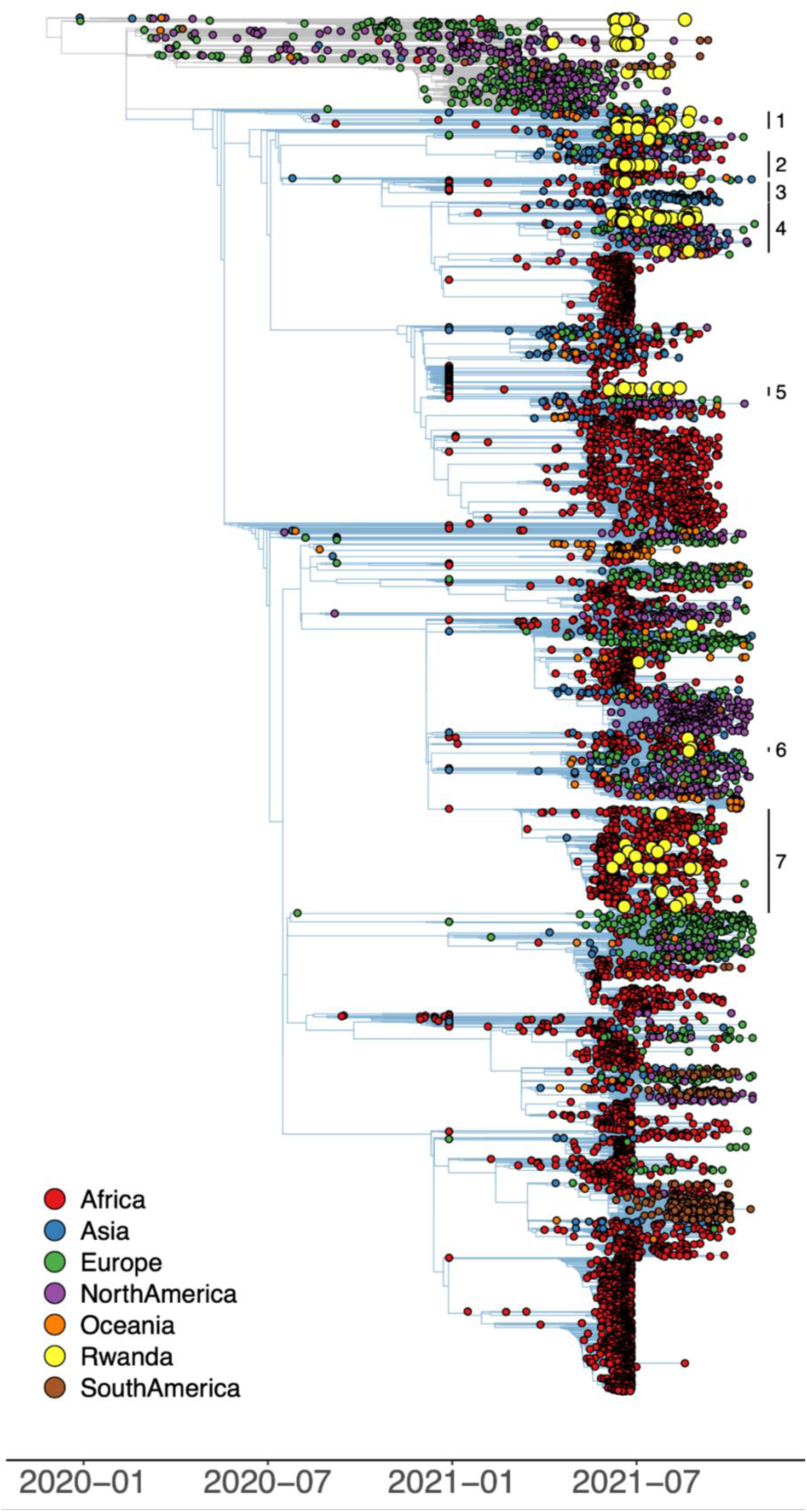
Time-calibrated phylogenetic tree of SARS-CoV-2 Delta (B.1.617.2 lineage) genome sequences. Branches connecting background genomes - including the non-Delta genomes generated in this study - are shown in grey, whereas branches connecting Delta genomes are shown in blue. Seven clusters of Rwandan Delta sequences can be visually identified, with other individual Rwandan sequences scattered throughout the overall Delta phylogeny.

Our global discrete phylogeographic analysis, based on a fixed phylogeny and including 315 Delta genomes collected in Rwanda (Figure 8), identified a minimum number of 66 distinct introduction events of the Delta variant (95% HPD interval = [64-68]) into Rwanda (Supplementary Figure S2). This further confirms our earlier findings regarding the relative importance of incoming travelers on establishing the waves of A.23.1 and B.1.380 infections in Rwanda earlier in the pandemic (Butera et al., 2021). Note that the size of the data set, owing to the magnitude of the Delta wave of infections, did not permit performing a full Bayesian discrete phylogeographic analysis between all locations while accommodating phylogenetic uncertainty.

Our follow-up continuous phylogeographic analysis within Rwanda consisted of a reconstruction along each Rwanda clade gathering at least three genomic sequences associated with geographic coordinates, and shows that introduction events into Rwanda did not lead to infectious clusters that remained constrained in limited geographic areas (Figure 9). In fact, we identified many long-range dispersal events that do not seem to involve the province of Kigali. Indeed, we estimate a total proportion of 0.20 (95% HPD = [0.12-0.28]) within-Kigali lineage dispersal events compared to 0.08 (95% HPD = [0.04-0.12]) and 0.05 (95% HPD = [0.03-0.07]) of lineage dispersal events exiting and arriving in Kigali (expressed in proportion of phylogenetic branches). We complement these estimates with the estimated proportions of lineage dispersal events occurring within a unique sector and between two different sectors. We find roughly as many inter-sectoral lineage dispersal events (0.44; 95% HPD = [0.38-0.52]) as intra-sectoral lineage dispersal events (0.54; 95% HPD = [0.43-0.61]), again expressed in proportion of phylogenetic branches.

**Figure 9.**
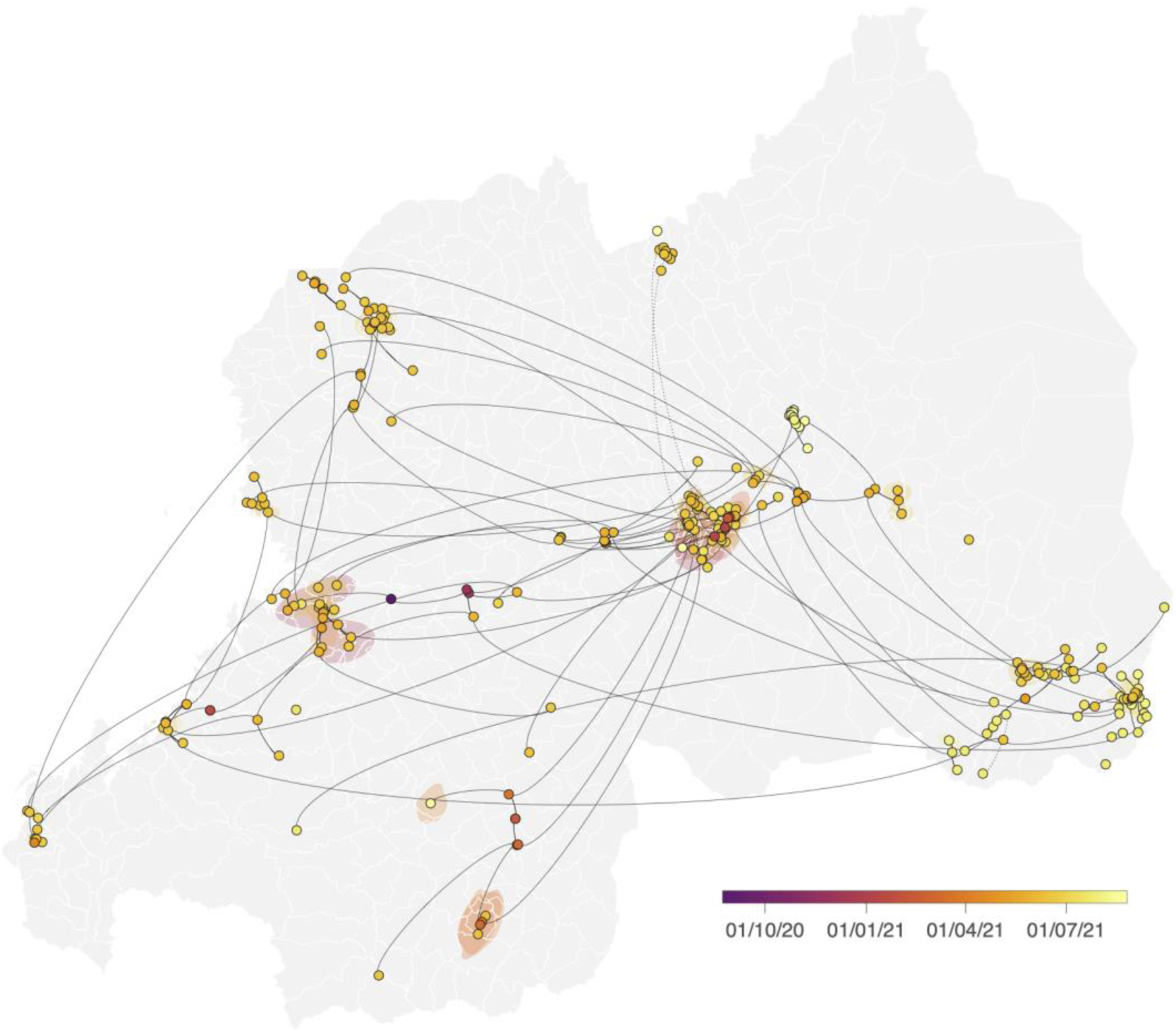
Spatially explicit phylogeographic reconstruction of the dispersal history of the Delta lineage in Rwanda. We performed continuous phylogeographic reconstruction along each Rwandan cluster identified by the initial discrete phylogeographic analysis (Figure S2). For each cluster, we mapped the maximum clade credibility (MCC) tree and overall 80% highest posterior density (HPD) regions reflecting the uncertainty related to the phylogeographic inference. MCC trees and 80% HPD regions are based on 1,000 trees subsampled from each post-burn-in posterior distribution. MCC tree nodes were colored according to their time of occurrence, andt 80% HPD regions were computed for successive time layers and then superimposed using the same color scale reflecting time. Continuous phylogeographic reconstructions were only performed along Rwandan clades linking at least three sampled genomic sequences generated in this study for which the geographic origin was known. Besides the phylogenetic branches of MCC trees obtained by continuous phylogeographic inference, we also mapped sampled sequences belonging to clades linking less than three geo-referenced sequences.

Among the 201 Rwandan Delta sequences in our dataset, 109 were associated with non-vaccinated patients and 92 with vaccinated patients. This represents a vaccination rate of 45% for our sample, which is not reflected in the overall rate in the country at the time, as at the beginning of September, the percentage of the population having received at least one dose was only 12%. We could speculate that this is due to the socio-economic status (SES) of the patients in the sample, as previous studies have shown an association between higher SES and access to healthcare (especially to primary care facilities). However, by far the most common occupation in our sample was farmer (*n* = 92), and among patients with this occupation, vaccination coverage was still 39%. Figure 2 shows that the vaccination coverage is higher in the capital region compared to the rest of the country, and we do have 37 sequences from Kigali in our dataset, but this alone does not explain such a high vaccination rate. Besides the slightly higher number of sequences from Kigali, we did not observe any clear geographical biases (such as a rural/urban divide) in our sample that might explain this discrepancy. Finally, our data set might exhibit unequal sampling of people living close to testing centers, and since these also often correspond to vaccination centers, it is possible that this explains why vaccination coverage in our data set is much higher compared to the national level of vaccination.

We identified 11 cases of breakthrough infection in vaccinated individuals in the period of more than two weeks after full vaccination, also known as breakthrough infections or (BTI), which corresponds to a breakthrough rate of 5.5% of total infections in our data set. Research suggests that Delta is associated with a higher rate of breakthrough infections compared to the initial Alpha strain, but less so compared to Omicron (Christensen et al., 2022; Goga et al., 2021). Studies have found breakthrough rates of 23% in Texas, USA (Christensen et al., 2022) and 32% in Singapore (Chia et al., 2022). Note that the percentage of vaccinated individuals plays a significant role in the rate of BTI, as in a highly vaccinated population we would expect higher rates of BTI.

## Discussion

In our study, we have performed representative sampling to the best of our abilities, by targeting samples from all provinces and different points of entry into the country, and hotspots that were mainly centered in the capital city of Kigali, and secondary towns. However, we do acknowledge a limitation in our study in the form of suboptimal sampling in the North-East of Rwanda, which is mostly covered by the Akagera National Park and where the border to Uganda had been closed from February 2019 to January 2022.

We can compare our own results to those of a recent study in Benin which focused on the emergence of the Delta lineage (Yadouleton et al., 2022). In Benin, Delta first appeared around May and became dominant by June 2021, similarly to what was observed in Rwanda and in some other neighbouring African countries (see Figure 6). Shortly after Delta became dominant (in August and September 2021), a spike in cases was observed, which is again similar to what was observed in our own study. A large COVID-19 wave around this time period was also observed in Malawi, Zambia, Kenya and Uganda. While Benin is far removed geographically from Rwanda, we also observe notable similarities when comparing the Rwandan lineage replacement patterns with those of Uganda, Rwanda’s neighboring country to the north. A recent Ugandan study using 266 naso-oropharyngeal samples collected during June-December 2021 shows the same pattern of replacement of A.23.1 by Delta, starting March 2021 and peaking in June 2021 (Bbosa et al., 2022). This similarity in patterns is not surprising as this was previously observed with A.23.1, when it became the dominant lineage in both countries by the end of 2020 (Figure 5, (Bugembe et al., 2021). Moreover, our previous study showed frequent mixing between lineages from Uganda and Rwanda through a travel history-aware Bayesian phylogeographic reconstruction (Butera et al., 2021). We speculate this similarity in patterns to continue in the future as lineages from both countries continue to mix, such as with the Omicron variant, where travel cases originating from Rwanda have already been observed in Uganda (Bbosa et al., 2022).

The geographic distribution of the collected genomes in this study shows Delta dominating the samples in the South-East, North-East, and central areas of Rwanda (Figure 4). Specifically, the presence of Delta samples close to the borders with Tanzania, the Democratic Republic of Congo and - to a lesser extent - Uganda makes it interesting to check for the presence of Delta genomes in those countries, in areas close to the border with Rwanda. Unfortunately, Tanzania has no publicly available Delta genomes whereas Burundi has 57 publicly available Delta genomes, but they lack precise geographic information other than that they were sampled in Burundi. A similar scenario holds for Uganda, from which 457 Delta genomes are publicly available, but without detailed sampling location information. Finally, the Democratic Republic of Congo has so far shared 501 Delta genomes, of which 183 came from Bukavu, a city close to the western border with Rwanda. While potentially interesting to link these samples to our phylogeographic analyses for Rwanda, they were all sampled within an 18-day interval, i.e., between June 13th and June 30th of 2021. This rather restricted temporal coverage only covered (part of) the first of four months of data collection in our study, and we have hence refrained from performing a phylogeographic analysis that could link these samples to those in Rwanda.

The majority of the population in Rwanda was vaccinated twice by the end of our study period. Specifically, 49.6% had been vaccinated with the Pfizer vaccine, 32.2% received the Moderna vaccine, whereas 12.8% received the AstraZeneca and 3.9% the Sinopharm vaccine. The vaccination coverage in the capital region is higher when compared to the rest of the country. This seems to point to there not being pro-rural inequality in terms of missed opportunities for vaccination in Rwanda, which were shown to be widespread across sub-Saharan Africa, driven mostly by household wealth (Adamu et al., 2019), instead, it might point to there being pro-urban inequality. While the sample size is relatively small, our data seemingly contains a disproportionately large number of breakthrough infections in patients vaccinated with AstraZeneca. Interestingly, in an analysis of more than 130 SARS-CoV-2-infected healthcare workers across three centers in India during a period of mixed lineage circulation, researchers observed reduced AstraZeneca vaccine effectiveness (nearly all had received two doses at least 21 days previously) against B.1.617.2 relative to non-B.1.617.2, with the caveat of possible residual confounding (Mlcochova et al., 2021). Additional data will have to be collected, and possibly also from the Omicron wave of infections, to make more conclusive statements on factors impacting BTIs in fully vaccinated individuals.

## Methods

### Study design

This is a follow-up study of SARS-CoV-2 strains that were circulating in Rwanda from June to September 2021, in which we describe the demography and epidemiology of 201 SARS-CoV-2 genomes from collected SARS-CoV-2-positive oropharyngeal swabs (Butera et al., 2021). These swabs were randomly selected from individuals residing in different provinces of Rwanda. There are 17 sites that conduct RT-PCR based testing for SARS-CoV-2, distributed across the four provinces of Rwanda, and the city of Kigali (Figure 1). There are five sites in Western province, four sites in Eastern province and in the capital City of Kigali, and two sites in the Northern and Southern provinces. These testing sites are located at teaching and referral hospitals in the country. Each teaching and referral hospital also has health centers within their catchment area where symptomatic patients go for sample collection, after which samples are sent to testing sites. Additionally, teaching and referral hospitals in collaboration with the Rwanda Biomedical Center (RBC) conduct regular mass screening campaigns countrywide (stadiums, schools, markets, bus stops). There is a SARS-CoV-2 testing site at Kigali International Airport, where all inbound travelers are tested upon arrival in Rwanda. Each testing site sends 10% of their samples to the National Reference Laboratory (NRL) monthly. All samples in this study were extracted from the biorepository of the NRL, in Kigali, Rwanda. Samples with a cycle threshold (*Ct*) value below 33 were selected, ensuring a wide geographical representation as well as ports of entry, and case description variables (date and place of RT-PCR test, age, sex, occupation, residence, nationality, travel history) were recorded.

### RNA Extraction

Ribonucleic acid (RNA) of the virus was extracted from confirmed SARS-CoV-2-positive clinical samples with *Ct* values ranging from 13.4 to 32.7 on a Maxwell 48 device using the Maxwell RSC viral RNA kit (Promega) following a viral inactivation step using proteinase K according to the manufacturer’s instructions.

### SARS-CoV-2 whole genome sequencing

Reverse transcription was performed using SuperScript IV VILO master mix, and 3.3 μl of RNA was combined with 1.2 μl of master mix and 1.5 μl of H2O. This was incubated at 25°C for 10 min, 50°C for 10 min, and 85°C for 5 min. PCRs used the primers and conditions recommended in the nCoV-2019 sequencing protocol (ARTIC Network, 2020) or the 1,200 bp amplicons described by Freed and colleagues ^21^. Primers from version 3 of the ARTIC Network and the 1,200 bp amplicons were used and were synthesized by Integrated DNA Technologies. Samples were multiplexed using the Oxford Nanopore native barcoding expansion kits 1-12, 13-24, or the native barcoding expansion 96 in combination with the ligation sequencing kit 109 (Oxford Nanopore). Sequencing was carried out on a MinION using R9.4.1 flow cells.

### Genome assembly

The data generated via the Oxford Nanopore Technology (ONT) MinION was processed using the ARTIC bioinformatic protocol (https://artic.network/ncov-2019/ncov2019-bioinformatics-sop.html). Briefly, the FAST5 sequence files were base called and demultiplexed using Guppy 4.2.2 in high accuracy mode, requiring barcodes at both ends of the read. FASTQ reads associated with each sample were filtered and concatenated via the guppy plex module. Consensus SARS-CoV-2 sequences were generated via the ARTIC nanopolish pipeline and assembled for each sample by aligning the respective sample reads to the Wuhan-Hu-1 reference genome (GenBank Accession: MN908947.3) with the removal of sequencing primers, followed by a polishing step using the raw Fast5 signal files. Positions with insufficient genome coverage, i.e. less than 30x, were masked with N.

### Phylogenetic analysis

Despite the public availability of an Africa-focused Nextstrain build (https://nextstrain.org/ncov/africa), the preferential sampling of more recently collected genomes (relative to the time of access) in such a build makes it so that attempting to place the Rwandan genomes generated in this study within this phylogenetic context results in a phylogeny unable to comprehensively capture the genetic diversity of SARS-CoV-2 circulating at the time of our study. Thus, we utilized the Nextstrain toolkit to perform a custom Africa-focused subsampling of SARS-CoV-2 genomes collected between December 31, 2019, and October 30, 2021. These sequences were further complemented to include all available Rwandan sequences on GISAID, as well as all available African Delta genomes collected prior to July 1, 2021. The cut-off date for these Delta genomes was selected to capture as much of the genetic diversity in the continent by the time Delta had become dominant in Rwanda. A preliminary analysis without these additional sequences showed a gap in the temporal coverage in our sampling, which in turn resulted in long branches in our downstream phylogenetic analysis. We aimed to potentially fill in this gap – i.e. split the long branches – by adding more sequences in order to include as many intermediary genomes that might have led to Rwandan introductions. This allowed us to more reliably identify major Rwandan Delta clades to use as a basis to study local circulation.

This workflow resulted in a total of 9,489 genomes considered for the phylogenetic analysis in this study. First, the 201 Rwandan SARS-CoV-2 genomes generated in this study were assigned Pango lineages, as described by Rambaut et al., using pangolin v2 and pangoLEARN model v2021-02-21 (O’Toole et al., 2021). We used Squarify to construct the square treemaps of lineage diversity across three time points (https://github.com/laserson/squarify). We mapped the combined data set against the canonical reference (GISAID ID: EPI_ISL_406801) using minimap2 and trimmed the data to positions 265-29,674 and padded with Ns in order to mask out 3’ and 5’ UTRs. We used the resulting alignment to estimate an unrooted maximum-likelihood phylogeny using IQ-TREE v2.1.2 under a general time-reversible model with empirical base frequencies and assuming among-site rate heterogeneity by means of a discretized gamma distribution with four rate categories (GTR+F+Γ4). We subsequently calibrated this phylogeny in time using TreeTime and the default Nextstrain settings – rooting the phylogeny on the Wuhan/Hu-1/2019 genome and fixing the clock rate to 0.0008 substitutions per site per year. We removed 101 outliers detected by TreeTime, which resulted in a time-calibrated phylogeny containing 9,397 taxa. We used ggtree (Yu et al., 2017) to visualize the resulting phylogeny.

### Phylogeographic analysis

We used the time-calibrated phylogeny from the previous section to perform spatially-explicit phylogeographic reconstruction of the dispersal history of the Delta lineage in Rwanda. We first performed a Bayesian discrete phylogeographic analysis (Lemey et al., 2009) on this phylogeny with only two (potential ancestral) location states (Rwanda/non-Rwanda) to identify internal nodes and descending clades that likely correspond to distinct introductions and ensuing clades (or “clusters”) into Rwanda (Dellicour et al., 2021). We ran this Bayesian inference through Markov chain Monte Carlo (MCMC) in BEAST 1.10 for 3×10^5^ generations and sampled every 1,000th iteration. We used the BEAGLE 3.2 high-performance computational library (Ayres et al., 2019) to increase computational efficiency. We inspected convergence and mixing aspects of all relevant parameters using Tracer 1.7 (Rambaut et al., 2018) to ensure that their associated effective sample size (ESS) values were all >200. After having discarded 10% of sampled posterior trees as burn-in, we constructed a maximum clade credibility (MCC) tree using TreeAnnotator 1.10 (Suchard et al., 2018). We used the resulting MCC tree to delineate Rwandan clades corresponding to independent introduction events (and corresponding 95% highest posterior density [HPD] interval) in Rwanda. In practice, we identified introduction events by comparing the locations assigned to each pair of nodes connected by the phylogenetic branches of the MCC tree, i.e. the most probable location inferred at internal nodes and the sampling location for tip nodes. We considered an introduction event to be the case when the location assigned to a node was “Rwanda” and the location assigned to its parent node in the tree was “non-Rwanda” (Dellicour et al., 2021).

We subsequently performed continuous phylogeographic reconstruction along each Rwandan cluster identified by the initial discrete phylogeographic analysis. We employed a relaxed random walk (RRW) diffusion model to obtain a posterior distribution of trees with estimated ancestral geographic coordinates for the internal nodes of the phylogeny, using a Cauchy distribution to model the among-branch heterogeneity in diffusion velocity (Lemey et al., 2010). We performed a distinct continuous phylogeographic reconstruction for each Rwandan clade identified by the initial discrete phylogeographic inference, again fixing a time-scaled subtree as an empirical tree, according to a previously published workflow (Dellicour et al., 2021). The clade-specific continuous phylogeographic reconstructions were only based on Rwandan tip nodes for which the Rwandan sector of origin was known and we only performed a continuous phylogeographic inference for Rwandan clades linking a minimum of three tip nodes with a known sector of origin. Sampling coordinates assigned to those genomic sequences were retrieved from a point randomly sampled within each sector of origin. Each Markov chain was run for 2×10^7^ generations and sampled every 10^5^ generations. MCMC convergence and mixing aspects were again assessed using Tracer 1.7 (Rambaut et al., 2018), and MCC trees (one per clade) were obtained with TreeAnnotator 1.10 (Suchard et al., 2018) after discarding 10% of the sampled trees as burn-in.

We used functions available in the R package “seraphim” (Dellicour et al., 2016) to extract spatiotemporal information embedded within the same 1,000 posterior trees and visualize the continuous phylogeographic reconstructions. For each cluster, we mapped the maximum clade credibility (MCC) tree and overall 80% highest posterior density (HPD) regions reflecting the uncertainty related to the phylogeographic inference. MCC trees and 80% HPD regions are based on 1,000 trees subsampled from each post-burn-in posterior distribution. MCC tree nodes were colored according to their time of occurrence, and 80% HPD regions were computed for successive time layers and then superimposed using the same color scale reflecting time. Besides the phylogenetic branches of MCC trees obtained by continuous phylogeographic inference, we also mapped sampled sequences belonging to clades linking less than three geo-referenced sequences.

### Estimating Delta’s growth rate advantage

To estimate the growth rate advantage between Delta and all other lineages of interest, we fitted a multinomial logistic spline model using all available Rwandan sequences in GISAID with complete collection date (n=682) using R version 3.6.3. Under this model, we estimated the additive increase in growth rate of Delta relative to other variants, as well as its transmission advantage (i.e. the expected percentage increase in effective reproductive number) (Campbell et al., 2021; Davies et al., 2021; Giovanetti et al., 2021). Given the variation in effective reproduction number across time, we averaged our estimates of transmission advantage across the time period between the collection dates of the first and last genomes of Delta sequenced in Rwanda. We used the *multinom* function of the *nnet* package to fit the model, and the *emtrends* function from the *emmeans* package to obtain the growth advantage estimates.

## Data Availability

All data produced in the present study are available upon reasonable request to the authors

## Ethical approval

The study was approved by the Rwanda National Ethics Committee (FWA Assurance No. 00001973 IRB 00001497 of IORG0001100/15April2020).

## Acknowledgements

We would like to acknowledge the work of Christian Happi, Enatha Mukantwari, Idowu Olawoye, Paul Oluniyi, Umuringa Jeanne D’arc, Esperance Umumararungu, Arlene Uwituze and Onikepe Folarin. This research was supported by the Academié de Recherche et Enseignement Supérieur (ARES) and from the Government of Rwanda through RBC/National Reference Laboratory in collaboration with the Belgian Development Agency (ENABEL) for additional genomic sequencing at GIGA Research Institute-Liege/Belgium. The views expressed in this publication are those of the authors and not necessarily those of the NIHR, the National Institute of Health Research, the Department of Health and Social Care or the Rwandan Government. SLH acknowledges support from the Research Foundation - Flanders (*Fonds voor Wetenschappelijk Onderzoek - Vlaanderen*, G0D5117N). NB, SD and GB acknowledge support from the Research Foundation - Flanders (*Fonds voor Wetenschappelijk Onderzoek - Vlaanderen*” G098321N). SD also acknowledges support from the *Fonds National de la Recherche Scientifique* (F.R.S.-FNRS, Bel-gium; grant n°F.4515.22) and from the European Union Horizon 2020 project MOOD (grant agreement n°874850). VH was supported by the Biotechnology and Biological Sciences Research Council (BBSRC) [grant number BB/M010996/1]. A.OT is supported by the Wellcome Trust Hosts, Pathogens & Global Health Programme [grant number: grant.203783/Z/16/Z] and Fast Grants [award number: 2236]. GB acknowledges support from the Internal Funds KU Leuven (Grant No. C14/18/094) and the Research Foundation - Flanders (“Fonds voor Wetenschappelijk Onderzoek - Vlaanderen,” G0E1420N). Computational resources have been provided by the *Consortium des Équipements de Calcul Intensif* (CÉCI), funded by the *Fonds de la Recherche Scientifique de Belgique* (F.R.S.-FNRS) under Grant n°2.5020.11 and by the Walloon Region.

---

**Table 1.** Descriptive statistics for the 201 Rwandan genomes collected in this study. Although our sample is relatively small, some general trends can be observed. Besides children, the lowest vaccination coverage is seen in adults between 36-50 years old. Older adults actually have a lower coverage rate in this sample, despite being the most at risk and hence prioritized for vaccination. Rwandan nationals seem to have a higher coverage compared to non-Rwandan nationals in the sample. There does not seem to be a large difference between sexes.

| Characteristics | Vaccination status |  |  |
| --- | --- | --- | --- |
|  | Yes<br>number (%) | Not or Unknown<br>number (%) | Total |
| **Nationality** |  |  |  |
| *Rwanda* | 84 (46.7) | 96 (53.3) | 180 |
| *Other* | 8 (38.1) | 13 (61.9) | 21 |
| **Sex** |  |  |  |
| *Female* | 47 (45.2) | 57 (54.8) | 104 |
| *Male* | 45 (46.4) | 52 (53.6) | 97 |
| **Age\*** |  |  |  |
| *< 18 yrs* | 3 (15.8) | 16 (84.2) | 19 |
| *18 - 35 yrs* | 44 (47.3) | 49 (52.7) | 93 |
| *36 - 50 yrs* | 32 (59.3) | 22 (40.7) | 54 |
| *51+ yrs* | 12 (36.4) | 21 (63.6) | 33 |
| **Occupation** |  |  |  |
| *Employed* | 78 (48.7) | 82 (51.3) | 160 |
| *Not employed* | 6 (31.58) | 13 (68.42) | 19 |
| *Student* | 8 (36.4) | 14 (63.6) | 22 |
| **Overall** | 92 (45.77) | 109 (54.23) | 201 |

\*2 missing age

**Table 2.** Descriptive statistics for the 11 cases of breakthrough Delta infections in our sample. In most cases the patient had received the AstraZeneca vaccine, even though a majority (49.6%) of the population has received the Pfizer vaccine.

| **Nationality** |  |
| --- | --- |
| *Rwanda* | 9 |
| *Other* | 2 |
| **Sex** |  |
| *Male* | 4 |
| *Female* | 7 |
| **Age** |  |
| *<18 yrs* | 0 |
| *18 - 35 yrs* | 6 |
| *36 - 50 yrs* | 5 |
| *50+ yrs* | 0 |
| **Occupation** |  |
| *Employed* | 4 |
| *Not Employed/Other* | 3 |
| *Student* | 4 |
| **Vaccination** |  |
| *Astrazeneca* | 9 |
| *Moderna* | 1 |
| *Pfizer* | 1 |

## Notes

### Competing Interest Statement

The authors have declared no competing interest.

