## Supplementary Figure 1 for "SARS-CoV-2 genomic surveillance in Rwanda: Introductions and local transmission of the B.1.617.2 (Delta) variant of concern": Supplementary Figure S1.pdf

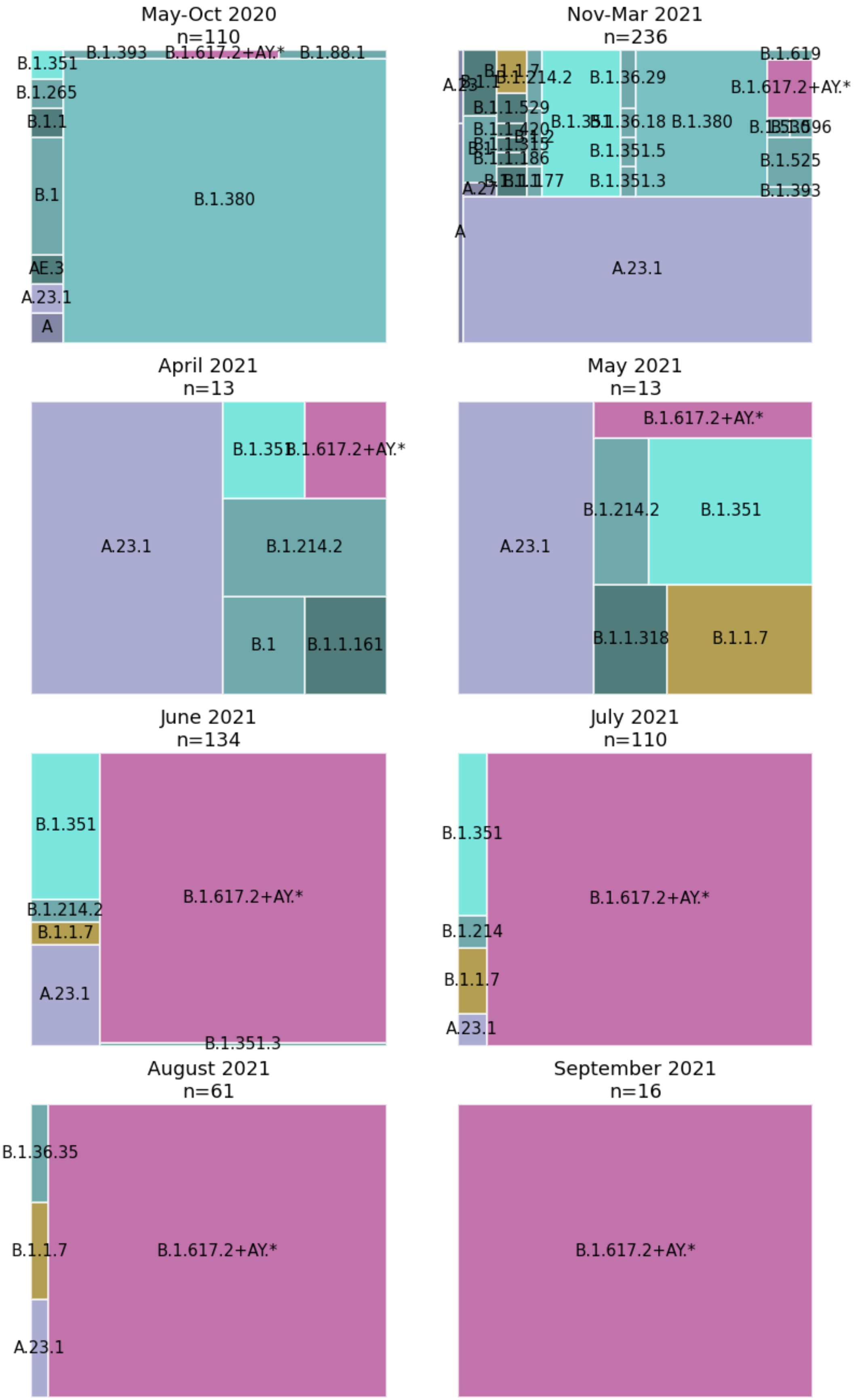
May-Oct 2020
n=110
B.1.351
B.1.393
B.1.617.2+AY.*
B.1.88.1
B.1.265
B.1.1
B.1
B.1.380
AE.3
A.23.1
A
Nov-Mar 2021
n=236
B.1.619
B.1.214.2
B.1.36.29
B.1.617.2+AY.*
B.1.1.529
B.1.36.18
B.1.380
B.1.525
B.1.1.186
B.1.351.5
B.1.351.3
B.1.393
A.27
A
A.23.1
April 2021
n=13
A.23.1
B.1.214.2
B.1
B.1.1.161
May 2021
n=13
B.1.617.2+AY.*
B.1.214.2
B.1.351
A.23.1
B.1.1.318
B.1.1.7
June 2021
n=134
B.1.351
B.1.617.2+AY.*
B.1.214.2
B.1.1.7
A.23.1
B.1.351.3
July 2021
n=110
B.1.351
B.1.617.2+AY.*
B.1.214
B.1.1.7
A.23.1
August 2021
n=61
B.1.36.35
B.1.1.7
B.1.617.2+AY.*
A.23.1
September 2021
n=16
B.1.617.2+AY.*

**Supplementary Figure S1.** Lineage diversity sampled in Rwanda across various time points, from the emergence of the B.1.380 and A.23.1 lineages in our previous study until the end of our current study period in September 2021. Lineage B.1.380, a Rwanda-specific lineage, dominated the sampled diversity during the first wave. Lineage A.23.1 first appeared in Rwanda in October 2020 and quickly attained a significant proportion of the sampled SARS-CoV-2 genome sequences. Lineage B.1.617.2 and its AY.* sublineages represent the majority of samples sequenced since June 2021, becoming completely dominant by the end of our study in September 2021.
