## Supplementary Figure 2 for "SARS-CoV-2 genomic surveillance in Rwanda: Introductions and local transmission of the B.1.617.2 (Delta) variant of concern": Supplementary Figure S2.pdf

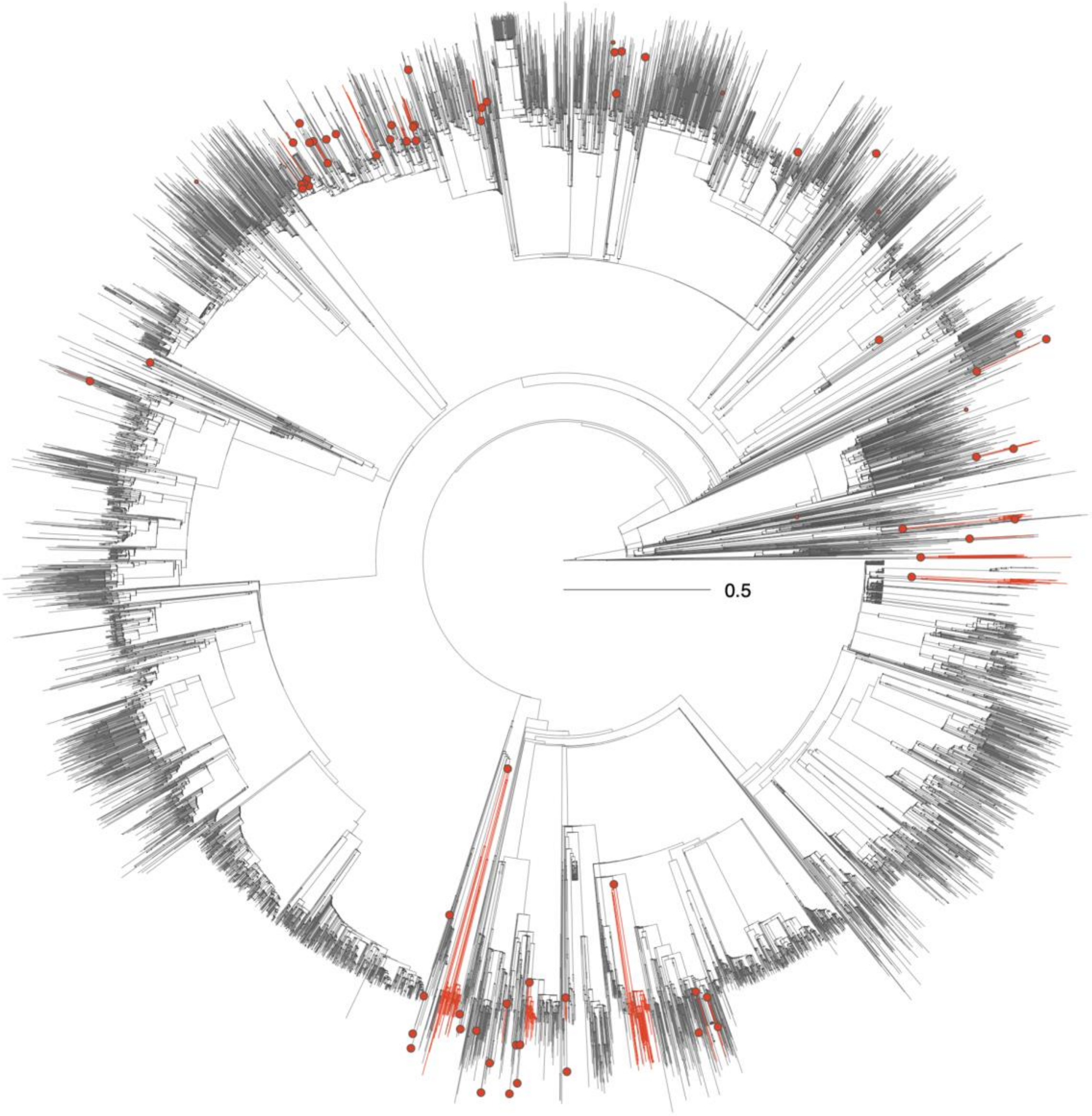


**Supplementary Figure S2.** Time-scaled phylogenetic tree in which we identified Rwandan clusters based on the genomes generated in this study. A cluster is here defined as a phylogenetic clade likely corresponding to a distinct introduction into Rwanda. We delineated these clusters by performing a discrete phylogeographic reconstruction along the time-scaled phylogenetic tree while only considering two potential ancestral locations: “Rwanda” and “non-Rwanda”. We identified a minimum number of 66 Delta lineage introductions (95% HPD interval = [64-68]) into Rwanda from our discrete phylogeographic analysis, which showcases the relative importance of external introductions considering the number of Delta sequences currently sampled in Rwanda. On the tree, lineages circulating in Rwanda are highlighted in red, and red nodes correspond to the most ancestral node of each Rwandan cluster.
