## Supplementary Table 1 for "SARS-CoV-2 genomic surveillance in Rwanda: Introductions and local transmission of the B.1.617.2 (Delta) variant of concern": Supplementary Table 1.pdf

| Date | Measures |
|---|---|
| 12 June 2021 | - Movements prohibited between 9 PM - 4 AM<br>- Public transport to not exceed 50% capacity; mask obligatory<br>- Arriving and departing passengers at Kigali International Airport to have a negative COVID-19 PCR test taken within 72 hours before departure<br>- Public offices, restaurants, and cafés operate at 30% capacity<br>- All swimming pools and spas are closed except for those at hotels hosting tested guests<br>- Social gatherings happening in homes are prohibited |
| 21 June 2021 | - Movements prohibited between 7 PM - 4 AM<br>- Movements between capital City of Kigali and other provinces are prohibited<br>- Physical meetings to not exceed 30% of the venue capacity and participants to present a negative SARS-CoV-2 test<br>- Public offices to operate at 15% capacity |
| 29 June 2021 | - Movements prohibited between 6 PM - 4 AM<br>- Public and private offices are closed<br>- Schools are closed<br>- Restaurants are closed and only offer take away services |
| 14 July 2021 | - Total lockdown in Kigali, Burera, Gicumbi, Kamonyi, Musanze, Nyagatare, Rubavu, Rwamagana, and Rutsiro<br>- Movements prohibited except for essential services like healthcare and groceries shopping<br>- Public transportation is stopped |
| 11 August 2021 | - Lockdown lifted<br>- Movements prohibited between 8 PM - 4 AM<br>- Public and private offices operate at 50% capacity<br>- Physical meetings to not exceed 30% of the venue capacity and participants to present a negative SARS-CoV-2 test<br>- Restaurants to operate at 30% capacity |

| | |
|---|---|
| 1 September 2021 | - Movements prohibited between 10 PM - 4 AM<br>- Public transport to operate at 75% capacity |

1.https://www.primature.gov.rw/fileadmin/user_upload/Primature/Publications/Cabinet_Decisions/English/_2021_Cabinet_Resolutions/Cabinet_Resolutions_of_12_June_2021___ENG_.pdf

2.https://www.primature.gov.rw/fileadmin/user_upload/Primature/Publications/Cabinet_Decisions/English/_2021_Cabinet_Resolutions/Cabinet_Communique_21_June_2021.pdf

3.https://www.primature.gov.rw/fileadmin/user_upload/Primature/Publications/Cabinet_Decisions/English/_2021_Cabinet_Resolutions/Communique.pdf

4.https://www.primature.gov.rw/fileadmin/user_upload/Primature/Publications/Cabinet_Decisions/English/_2021_Cabinet_Resolutions/Cabinet_communique_July_14th__2021.pdf

5.https://www.primature.gov.rw/fileadmin/user_upload/Primature/Publications/Cabinet_Decisions/English/_2021_Cabinet_Resolutions/Cabinet_Communique__11th_August_2021.pdf

6.https://www.primature.gov.rw/fileadmin/user_upload/Primature/Publications/Cabinet_Decisions/English/_2021_Cabinet_Resolutions/Cabinet_Resolutions_of_1st_Sept_2021__ENG_.pdf

**Supplementary Table S1.** Non-pharmaceutical interventions by announcement date in Rwanda during our study period, with key changes described. Sources are official government communiques, provided beneath the table.
